# Underlying causes of death with changing mortality among adults in the United States, 2013-2017

**DOI:** 10.1101/2020.06.13.20130450

**Authors:** Xin Hu, Yong Lin, Gangjian Qin, Lanjing Zhang

## Abstract

**Background and objectives:** Overall mortality among U.S. adults was stable in the past years, while racial disparity was found in 10 leading causes of death or age-specific mortality in Blacks or African Americans. However, the trends in sex- and race-adjusted age-standardized cause-specific mortality are poorly understood.

**Methods:** We here aimed at identifying the UCD with sex- and race-adjusted, age-standardized mortality that was changing in recent years. We extracted the data of underlying causes of death (UCD) from the Multiple Cause of Death database of the Centers for Disease Control and Prevention (CDC). Multivariable log-linear regression models were used to estimate trends in sex- and race-adjusted, age-standardized mortality of UCD during 2013-2017.

**Results:** A total of 31,029,133 deaths were identified. Among the list of 113 UCD compiled by the CDC, there were 29 UCD with upward trend, 33 UCD with downward trend and 56 UCD with no significant trend-changes. The 2 UCD with largest annual percent change were both nutrition related (annual percent change= 17.73, 95% CI [15.13-20.33] for malnutrition, and annual percent change= 17.49, 95% CI [14.94-20.04] for Nutritional deficiencies), followed by Accidental poisoning and exposure to noxious substances.

**Conclusions:** This study thus reported the UCD with changing mortality in recent years, which was sex- and race-adjusted and age-standardized. More efforts and resources should be focused on understanding, prevention and control of the mortality linked to these UCD. Continuous monitoring of mortality trends is recommended.

---

Overall mortality among U.S. adults was stable in the past years.^1 2^ Recent works documented the racial disparity in 10 leading causes of death or age-specific mortality in U.S. Blacks or African Americans.^3 4^ and disparities of race, age, and sex in the trends of suicide mortality.^5^ However, the trends in sex- and race-adjusted age-standardized cause-specific mortality are largely unknown, despite its significance in public health, policy making and disease prevention. We therefore aimed to describe the Underlying causes of death (UCD) with sex- and race-adjusted, age-standardized mortality that was changing in recent years (2013-2017).

## Methods

Multiple Cause of Death database of the Centers for Disease Control and Prevention (CDC) contains mortality and population counts for all U.S. counties.^6^ Its data were extracted from death certificates for all eligible U.S. residents. The age-standardized mortality rates during 2013-2017 were estimated for adults (25+ years) in the U.S., using the U.S. standard population of year 2000 and the CDC Multiple Cause of Death database (1999-2017).^6^ UCD were the single, underlying cause of death reported on each death certificate, and here classified using the list of 113 UCD compiled by the CDC.^1 2^ Age-standardized mortality rates by sex and race (White versus Non-white) and multivariable log-linear regression models were used for computing annual percent change (APC) of sex- and race-adjusted, age-standardized mortality rates, according to the CDC guidelines on using NHCS data.^7^ The Stata software (version 15, StataCorp, College Station, TX) was employed for statistical analyses. An Institutional Review Board (IRB) approval is not required due to the use of de-identified and publicly available data on the deceased subjects. All *P* values were 2-sided and considered significant if <.05. The trends during 2015-2017 were also computed to assess potential influence of implementation of the 10th edition of International Classification of Diseases (ICD-10) in 2015.

## Results

Among 31,027,401 deaths with un-suppressed mortality rate recorded during 2013-2017 (99.9% of the total deaths), there were 14,814,842 women (47.8%) and 26,499,625 Whites (85.4%) (**Table 1**). In the listed 113 UCD, 29 UCD had an upward trend in the sex- and race-adjusted age-standardized morality during 2013-2017 (**Table 2** and **Figure 1**), including in the ranking order of APC: Malnutrition/nutritional deficiencies, Accidental poisoning and exposure to noxious substances, Other nutritional deficiencies, Complications of medical and surgical care, Hypertensive heart and renal disease, Acute and rapidly progressive nephritic and nephrotic syndrome, Other and unspecified events of undetermined intent and their sequelae, Nontransport accidents, Alzheimer disease, Events of undetermined intent, Accidents (unintentional injuries), Parkinson disease, Alcoholic liver disease and others. In these 29 UCD, the 2 UCD with largest APC were both nutrition related, followed by Accidental poisoning and exposure to noxious substances. A total of 7 accident-related UCD, 4 heart-related UCD, 3 hypertension-related UCD and 2 liver-related UCD had increasing sex- and race-adjusted age-standardized mortality. There were also 33 UCD with downward trend in sex- and race-adjusted age-standardized mortality (**Table 3**), including mostly infection, malignancies and ischemic-heart diseases. There were 56 UCD with no significant trend, and 15 UCD with statistically-suppressed morality rate. The multivariable sensitivity analysis identified 16 UCD with upward mortality trend and 13 UCD with downward mortality trend during 2015-2017 (data not shown), which all had similar trends during 2013-2017.

**Table 1.**
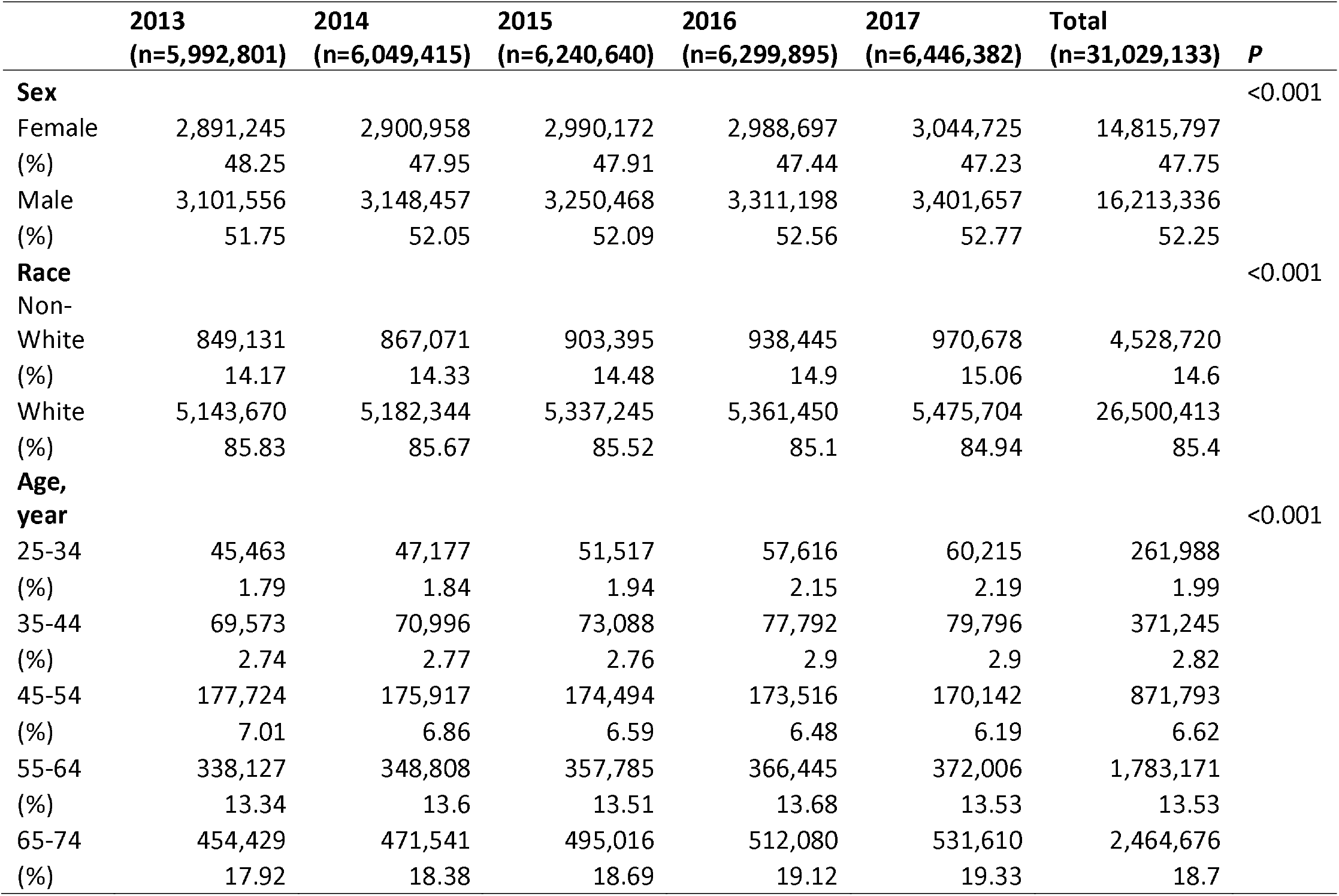
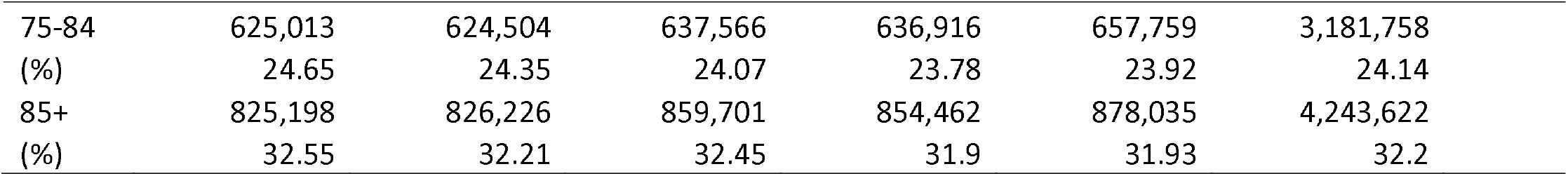
Baseline characteristics of deaths among adults in the U.S. by year, 2013-2017.

**Table 2.** The underlying causes of death with increasing sex- and race-adjusted age-standardized mortality among adults in the U.S., 2013-2017.

| Underlying cause of death<sup>¶</sup> | 2013<br>mortality* | 2017<br>mortality* | Death, n | APC (95% CI) | P<sup>§</sup> |
| --- | --- | --- | --- | --- | --- |
| Malnutrition (E40-E46) | 1.37 | 2.95 | 25806 | 17.73 (15.13 to 20.33) | <0.001 |
| #Nutritional deficiencies (E40-E64) | 1.43 | 3.06 | 26788 | 17.49 (14.94 to 20.04) | <0.001 |
| Accidental poisoning and exposure to noxious substances (X40-X49) | 17.28 | 28.54 | 230327 | 13.89 (10.94 to 16.84) | <0.001 |
| #Complications of medical and surgical care (Y40-Y84,Y88) | 1.18 | 1.75 | 15203 | 10.36 (6.36 to 14.35) | <0.001 |
| Hypertensive heart and renal disease (I13) | 1.75 | 2.86 | 26402 | 10.01 (6.98 to 13.04) | <0.001 |
| Acute and rapidly progressive nephritic and nephrotic syndrome (N00–N01,N04) | 0.16 | 0.23 | 2550 | 8.96 (5.03 to 12.89) | <0.001 |
| Other and unspecified events of undetermined intent and their sequelae (Y10–Y21,Y25–Y34,Y87.2,Y89.9) | 1.79 | 2.25 | 21213 | 7.63 (2.31 to 12.95) | 0.011 |
| Nontransport accidents (W00–X59,Y86) | 39.14 | 52.04 | 501465 | 7.59 (5.44 to 9.74) | <0.001 |
| #Alzheimer disease (G30) | 36.38 | 48.01 | 526372 | 7.52 (6.13 to 8.91) | <0.001 |
| Events of undetermined intent (Y10–Y34,Y87.2,Y89.9) | 1.91 | 2.34 | 22257 | 7.44 (2.25 to 12.64) | 0.011 |
| #Accidents (unintentional injuries) (V01–X59,Y85–Y86) | 53.08 | 67.47 | 660823 | 6.57 (4.54 to 8.59) | <0.001 |
| #Parkinson disease (G20–G21) | 11.26 | 13.01 | 140968 | 4.67 (3.92 to 5.41) | <0.001 |
| Alcoholic liver disease (K70) | 7.88 | 9.17 | 102510 | 3.97 (2.26 to 5.68) | <0.001 |
| Motor vehicle accidents (V02–V04,V09.0,V09.2,V12–V14,V19.0–V19.2,V19.4..) | 12.91 | 14.39 | 147771 | 3.93 (2.59 to 5.26) | <0.001 |
| Transport accidents (V01–V99,Y85) | 13.93 | 15.44 | 159358 | 3.79 (2.56 to 5.02) | <0.001 |
| Heart failure (I50) | 27.80 | 31.56 | 367490 | 3.54 (2.95 to 4.13) | <0.001 |
| Hypertensive heart disease (I11) | 15.86 | 18.78 | 208511 | 2.96 (1.53 to 4.39) | 0.001 |
| Intentional self-harm (suicide) by other and unspecified means and their sequelae (*U03,X60–X71,X75–X84,Y87.0) | 8.16 | 9.15 | 92679 | 2.91 (1.35 to 4.47) | 0.002 |
| #Intentional self-harm (suicide) (*U03,X60–X84,Y87.0) | 16.89 | 18.27 | 190649 | 2.68 (1.66 to 3.70) | <0.001 |
| Other and unspecified infectious and parasitic diseases and their sequelae (A00,A05,A20–A36,A42–A44,A48–A49,A54–A79,A81–A82,A85.0–A85.1,A85.8,A86–B04,B06–B09,B25–B49,B55–B99) | 2.49 | 2.90 | 31630 | 2.66 (0.70 to 4.63) | 0.015 |
| Water, air and space, and other and unspecified transport accidents and their sequelae (V90–V99,Y85) | 0.65 | 0.67 | 7376 | 2.55 (0.39 to 4.70) | 0.032 |
| Malignant neoplasms of corpus uteri and uterus, part unspecified (C54–C55) | 3.97 | 4.28 | 50862 | 1.91 (1.42 to 2.39) | <0.001 |
| Other complications of pregnancy, childbirth and the puerperium (O10–O99) | 0.49 | 0.54 | 4881 | 1.87 (0.41 to 3.33) | 0.033 |
| #Pregnancy, childbirth and the puerperium (O00–O99) | 0.49 | 0.54 | 4976 | 1.73 (0.24 to 3.23) | 0.049 |
| Other diseases of respiratory system (J00–J06,J30–J39,J67,J70–J98) | 15.11 | 16.42 | 187819 | 1.71 (0.75 to 2.67) | 0.003 |
| Other and unspecified nontransport accidents and their sequelae (W20–W31,W35–W64,W75–W99,X10–X39,X50–X59,Y86) | 6.57 | 6.85 | 80381 | 1.64 (0.67 to 2.62) | 0.004 |
| #Essential hypertension and hypertensive renal disease (I10,I12,I15) | 13.17 | 13.92 | 161640 | 1.41 (0.24 to 2.57) | 0.029 |
| #Chronic liver disease and cirrhosis (K70,K73–K74) | 15.68 | 16.80 | 197021 | 1.38 (0.37 to 2.38) | 0.015 |
| Other heart diseases (I26–I51) | 84.05 | 87.94 | 1052221 | 0.91 (0.34 to 1.47) | 0.005 |

Note: <sup>¶</sup>The list of 113 underlying causes of death compiled by the U.S. Centers for Disease Control and Prevention, with the 10<sup>th</sup> edition of the International classification of Diseases (ICD-10) in parentheses; \*Age-standardized mortality per 100,000 in the U.S.; APC, annual percent change; CI, confidence intervals; <sup>¶</sup>P for sex- and race-adjusted trend in the mortality which was age-standardized using the U.S. standard population of year 2000; # indicates a "rankable" cause of death, from the National Center for Health Statistics (NCHS) list of rankable causes of death. The rankable causes are a subset of the 113 selected causes of death.

**Table 3.** The underlying causes of death with decreasing sex- and race-adjusted age-standardized mortality among adults in the U.S., 2013-2017.

| Underlying cause of death <sup>¶</sup> | 2013 mortality* | 2017 mortality* | Death, n | APC | (95% CI) | P<sup>¶</sup> |
| --- | --- | --- | --- | --- | --- | --- |
| #Viral hepatitis (B15–B19) | 3.31 | 2.13 | 35696 | -11.71 | (-13.31 to -10.11) | <0.001 |
| Bronchitis, chronic and unspecified (J40–J42) | 0.26 | 0.20 | 2672 | -8.26 | (-13.57 to -2.95) | 0.007 |
| Emphysema (J43) | 3.59 | 2.78 | 37036 | -7.11 | (-12.14 to -2.07) | 0.012 |
| #Human immunodeficiency virus (HIV) disease (B20–B24) | 3.17 | 2.43 | 31524 | -7.10 | (-13.48 to -0.72) | 0.042 |
| #Atherosclerosis (I70) | 2.83 | 2.17 | 30369 | -6.55 | (-7.75 to -5.34) | <0.001 |
| #Enterocolitis due to Clostridium difficile (A04.7) | 3.32 | 2.41 | 35022 | -6.47 | (-8.6 to -4.33) | <0.001 |
| Hodgkin disease (C81) | 0.48 | 0.39 | 5133 | -5.69 | (-9.28 to -2.09) | 0.006 |
| Certain other intestinal infections (A04,A07-A09) | 4.49 | 3.41 | 47250 | -5.55 | (-7.38 to -3.72) | <0.001 |
| Respiratory tuberculosis (A16) | 0.16 | 0.14 | 1808 | -5.27 | (-10.13 to -0.4) | 0.047 |
| Malignant neoplasms of trachea, bronchus and lung (C33-C34) | 67.06 | 56.63 | 760393 | -4.68 | (-6.78 to -2.57) | <0.001 |
| Symptoms, signs and abnormal clinical and laboratory findings, not elsewhere classified (R00-R99) | 14.68 | 11.74 | 149792 | -4.13 | (-6.39 to -1.88) | 0.002 |
| Pneumonia (J12-J18) | 22.72 | 19.31 | 251383 | -4.07 | (-5.39 to -2.75) | <0.001 |
| Acute myocardial infarction (I21-I22) | 50.11 | 43.36 | 566631 | -3.94 | (-5.29 to -2.58) | <0.001 |
| #Influenza and pneumonia (J09-J18) | 24.25 | 21.81 | 273633 | -3.27 | (-4.8 to -1.74) | <0.001 |
| Ischemic heart diseases (I20-I25) | 158.67 | 143.56 | 1830094 | -2.83 | (-3.93 to -1.73) | <0.001 |
| All other forms of chronic ischemic heart disease (I20,I25.1-I25.9) | 81.63 | 73.80 | 937255 | -2.82 | (-4.27 to -1.37) | 0.001 |
| Malignant neoplasm of stomach (C16) | 4.91 | 4.44 | 56378 | -2.58 | (-3.48 to -1.68) | <0.001 |
| Other forms of chronic ischemic heart disease (I20,I25) | 106.84 | 98.48 | 1242985 | -2.39 | (-3.4 to -1.38) | <0.001 |
| Leukemia (C91-C95) | 10.02 | 9.14 | 112661 | -2.25 | (-3.41 to -1.1) | 0.001 |
| Malignant neoplasms of kidney and renal pelvis (C64-C65) | 5.94 | 5.44 | 69758 | -2.22 | (-3.14 to -1.31) | <0.001 |
| Malignant neoplasms of lymphoid, hematopoietic and related tissue (C81-C96) | 24.60 | 22.79 | 282027 | -2.2 | (-3.25 to -1.15) | 0.001 |
| Non-Hodgkin lymphoma (C82-C85) | 8.77 | 8.13 | 100796 | -2.2 | (-2.99 to -1.41) | <0.001 |
| #Aortic aneurysm and dissection (I71) | 4.31 | 4.00 | 49207 | -2.14 | (-3.29 to -0.98) | 0.002 |
| #Malignant neoplasms (C00-C97) | 250.98 | 234.46 | 2955924 | -2.01 | (-2.4 to -1.63) | <0.001 |
| Malignant neoplasms of colon, rectum and anus (C18-C21) | 22.55 | 21.19 | 264027 | -1.77 | (-2.44 to -1.11) | <0.001 |
| Malignant neoplasm of ovary (C56) | 6.11 | 5.57 | 70677 | -1.67 | (-2.39 to -0.95) | 0.001 |
| Other diseases of circulatory system (I71-I78) | 8.22 | 7.80 | 96630 | -1.59 | (-2.48 to -0.7) | 0.002 |
| All other diseases (Residual) | 136.30 | 131.52 | 1599086 | -1.30 | (-2.07 to -0.54) | 0.004 |
| Other chronic liver disease and cirrhosis (K73-K74) | 7.80 | 7.68 | 94511 | -1.26 | (-2.39 to -0.13) | 0.042 |
| #Septicemia (A40-A41) | 16.39 | 16.13 | 197318 | -1.15 | (-1.89 to -0.41) | 0.007 |
| Atherosclerotic cardiovascular disease, so described (I25.0) | 25.26 | 24.65 | 305730 | -1.12 | (-1.83 to -0.42) | 0.006 |
| #Diseases of heart (I00-I09,I11,I13,I20-I51) | 261.74 | 254.45 | 3133923 | -1.01 | (-1.56 to -0.45) | 0.002 |
| Major cardiovascular diseases (I00-I78) | 341.71 | 336.23 | 4111472 | -0.60 | (-0.97 to -0.23) | 0.005 |

Note: <sup>¶</sup> The list of 113 underlying causes of death compiled by the U.S. Centers for Disease Control and Prevention, with the 10<sup>th</sup> edition of the International classification of Diseases (ICD-10) in parentheses; \*Age-standardized mortality per 100,000 in the U.S.; APC, annual percent change; CI, confidence intervals; <sup>¶¶</sup> P for sex- and race-adjusted trend in the mortality which was age-standardized using the U.S. standard population of year 2000; # indicates a "rankable" cause of
death, from the National Center for Health Statistics (NCHS) list of rankable causes of death. The rankable causes are a subset of the 113 selected causes of death.

**Figure 1.**
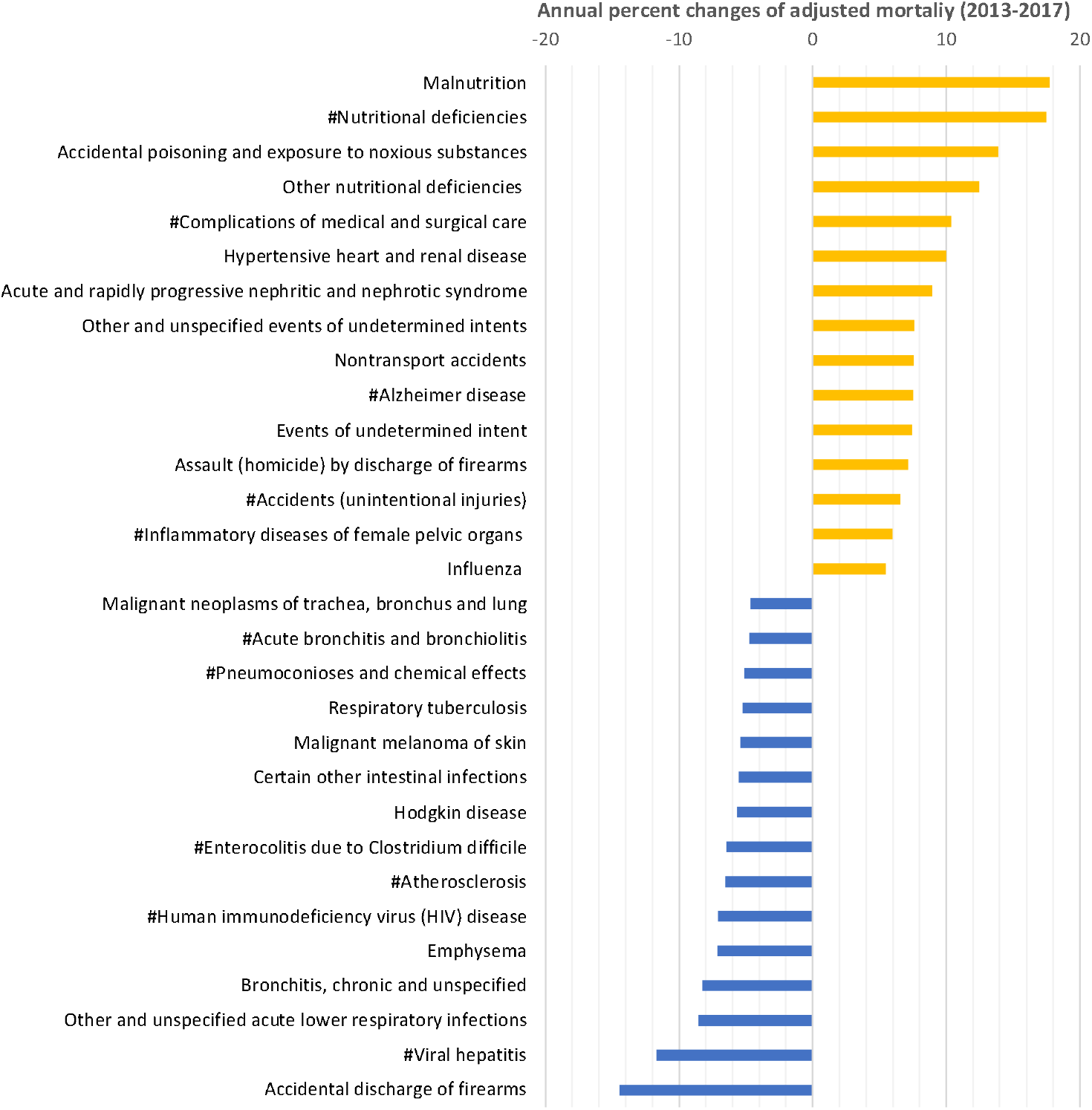
The top 15 underlying causes of deaths of the highest increasing and decreasing trends in sex- and race-adjusted age-standardized mortality among adults in the U.S., 2013-2017. The underlying causes of deaths with increasing trends were highlighted in yellow, while those with decreasing trends in blue. # indicates a “rankable” cause of death, from the National Center for Health Statistics list of rankable causes of death. The rankable causes are a subset of the 113 selected causes of death.

## Discussion

We here report the UCD with growing mortality and those with decreasing mortality among the adults in the U.S. during 2013-2017, which was sex- and race-adjusted and age-standardized. Sex- and race-adjustment reduces the biases associated with these disparity factors in mortality.^4 5 8^ The 2 UCD with largest APC were both nutrition related (APC= 17.73 for Malnutrition and APC= 17.49 for Nutritional deficiencies), followed by Accidental poisoning and exposure to noxious substances. Future policy making and public health resources perhaps should be focused on the identified UCD with growing mortality.

Despite the overall downward trends, cardiovascular diseases, heart diseases, and stroke mortality all had a decelerated downward trend substantially during 2011-2014.^9^ It is therefore possible, but not certain, that some of these UCD might be linked to increasing mortality in recent years. Moreover, trend analyses on mortality were rightfully focused on leading causes of death.^3 9 10^ Few studies systematically delineated recent trends of UCD with increasing mortality. It is probably most effective to decrease overall mortality by identifying and then reversing or neutralizing the increasing morality of certain UCD. Finally, age-standardized mortality had significantly different trends from those of crude mortality.^8^ Given the variations of morality trends by race, sex and age and the lack of related data, this study is important for reporting understand the sex- and race-adjusted age-standardized mortality, and identifying the UCD linked to changing mortality. It thus fills the knowledge gap of recent trends in sex- and race-adjusted age-standardized mortality, and provides early data for policy making and public health focus area on projecting and reducing future mortality among adults in the U.S.

Malnutrition/nutrition deficiency was one of the top-10 leading UCD among pediatric populations of American Indians/Alaska natives or Asian/Pacific Islanders in 2013,^1^ and Hispanic females aged 5-9 years in 2016.^2^ Given its ∼17.5% of APC, our data suggest malnutrition/nutrition deficiency will likely become a leading UCD in adults in the near future. The increasing mortality linked to UCD of accidents, heart, hypertension and liver are also concerning. Hypertension, for example, was among top-10 leading UCD in U.S. adults aged >85 years in 2016, but not in 2013.^1 2^ Future studies should continue monitoring UCD-specific mortality, and better understand and control these upward trends.

Several limitations of this study are noteworthy. First, the UCD on some death certificates may be misclassified or missed. For example, coders more likely include the UCD which the patient recently presented than the ones remotely presented. Second, there was a transition of ICD-9 to ICD-10 coding in 2015. Some cases may have different ICD-9 and ICD-10 classifications, but studies have shown that ICD-9 and ICD-10 classifications are overall consistent with each other.^11 12^ In addition, the list of 113 UCD were unlikely influenced by such a transition because of its relatively broad definition of each UCD (none of them had decimals). Finally, there were missing/suppressed data in some UCD due to the small number of deaths linked to them, which were excluded in the analysis.

In summary, this study reports the UCD that are associated with upward or downward trend in sex- and race-adjusted, age-standardized U.S. mortality rate during 2013-2017. Some of them are reported recently and consistent with our findings.^10^ These data will help better prioritize efforts on reducing mortality among U.S. adults.

### Future directions

Continuous monitoring of mortality trends is warranted. Future works should be focused on the causes of the rapidly increasing trends in certain UCD. Probably more importantly, we may consider the approaches to effectively monitor and prevent the further increases in deaths associated with these UCD.

## Data Availability

The data are available at CDC WONDER website.

## Abbreviations

APC: annual percent change
CDC: Centers for Disease Control and Prevention
ICD-10: the 10th edition of International Classification of Diseases
UCD: Underlying causes of death

## Declarations

### Authors’ contributions

LZ had full access to all of the data in the study and takes responsibility for the integrity of the data and the accuracy of the data analysis. Concept and design (LZ, GQ); drafting of the manuscript (XH, LZ); statistical analysis (XH, YL, LZ); supervision (LZ); acquisition, analysis, or interpretation of data (all authors); critical revision of the manuscript for important intellectual content (all authors).

## Bibliography

1. Heron MP. Deaths: leading causes for 2013. National vital statistics reports 2016;65(2) PMID:

2. Heron MP. Deaths: Leading causes for 2016. National vital statistics reports 2018;67(6) PMID:

3. Cunningham TJ, Croft JB, Liu Y, Lu H, Eke PI, Giles WH. Vital Signs: Racial Disparities in Age-Specific Mortality Among Blacks or African Americans - United States, 1999-2015. MMWR Morbidity and mortality weekly report 2017;66(17):444–56. doi: 10.15585/mmwr.mm6617e1, PMID: 28472021

4. Chang MH, Moonesinghe R, Athar HM, Truman BI. Trends in Disparity by Sex and Race/Ethnicity for the Leading Causes of Death in the United States-1999-2010. Journal of public health management and practice : JPHMP 2016;22 Suppl 1:S13–24. doi: 10.1097/phh.0000000000000267, PMID: b25946701

5. Ivey-Stephenson AZ, Crosby AE, Jack SPD, Haileyesus T, Kresnow-Sedacca MJ. Suicide Trends Among and Within Urbanization Levels by Sex, Race/Ethnicity, Age Group, and Mechanism of Death - United States, 2001-2015. Morbidity and mortality weekly report Surveillance summaries (Washington, DC : 2002) 2017;66(18):1–16. doi: 10.15585/mmwr.ss6618a1, PMID: 28981481

6. Centers for Disease Control and Prevention (CDC) NCfHSN. Multiple Cause of Death 1999-2017 on CDC WONDER Online Database, released 2018. Data are compiled from data provided by the 57 vital statistics jurisdictions through the Vital Statistics Cooperative Program.: U.S. HHS; 2018 [updated Feb 22, 2019. Available from: http://wonder.cdc.gov/mcd-icd10.html accessed June 18 2019.

7. Ingram DD, Malec DJ, Makuc DM, Kruszon-Moran D, Gindi RM, Albert M, et al. National Center for Health Statistics Guidelines for Analysis of Trends. Vital Health Stat 2 2018(179):1-71. PMID: 29775435

8. Murphy SL, Xu J, Kochanek KD. Deaths: final data for 2010. Natl Vital Stat Rep 2013;61(4):1-117. PMID: 24979972

9. Sidney S, Quesenberry CP, Jr., Jaffe MG, Sorel M, Nguyen-Huynh MN, Kushi LH, et al. Recent Trends in Cardiovascular Mortality in the United States and Public Health Goals. JAMA cardiology 2016;1(5):594–9. doi: 10.1001/jamacardio.2016.1326, PMID: 27438477

10. Tapper EB, Parikh ND. Mortality due to cirrhosis and liver cancer in the United States, 1999-2016: observational study. BMJ (Clinical research ed) 2018;362:k2817. doi: 10.1136/bmj.k2817, PMID: 30021785

11. Quan H, Li B, Saunders LD, Parsons GA, Nilsson CI, Alibhai A, et al. Assessing validity of ICD-9-CM and ICD-10 administrative data in recording clinical conditions in a unique dually coded database. Health Serv Res 2008;43(4):1424–41. doi: 10.1111/j.1475-6773.2007.00822.x, PMID: 18756617

12. Anderson RN, Minino AM, Hoyert DL, Rosenberg HM. Comparability of cause of death between ICD-9 and ICD-10: preliminary estimates. Natl Vital Stat Rep 2001;49(2):1-32. PMID: 11381674

